# LONG TERM PSYCHOLOGICAL CONSEQUENCES OF STROKE (OX-CHRONIC): A LONGITUDINAL STUDY OF COGNITION IN RELATION TO MOOD AND FATIGUE AFTER STROKE: PROTOCOL

**DOI:** 10.1101/2021.06.18.21259013

**Authors:** Nele Demeyere, Owen A Williams, Elise Milosevich, Evangeline Grace Chiu, Bogna Drozdowska, Avril Dillon, Helen Dawes, Shirley Thomas, Annapoorna Kuppuswamy, Sarah T Pendlebury, Terry Quinn

## Abstract

**Background:** The long-term psychological consequences of stroke and how cognitive problems change over time after the first-year following stroke remain unclear. Particularly, trajectories of domain-specific and domain-general cognitive functions and how cognition interacts with mood, fatigue, and quality of life are not well described.

**Aims:** To determine the prevalence, trajectories and wider impact of domain specific cognitive impairment in long term stroke survivors, in relation to mood, fatigue, and quality of life.

**Methods:** Participants who previously took part in the Oxford Cognitive Screening study, completed the 6-month follow up with cognitive, mood, fatigue, and quality of life assessments and agreed to be contacted for future research will be recruited into OX-CHRONIC. The eligible cohort is between 2- and 9-years post stroke. Cognition will be assessed with a detailed neuropsychological battery, alongside questionnaire measures of mood, fatigue, activities of daily life and quality of life measures at two timepoints, one year apart. Additionally, medical records will be accessed to extract further clinical information about the stroke and patients may opt-in to wear an activity monitor for one week to provide fine-grained measures of sleep and activity. The study protocol and study materials were approved by the national ethics committee (REC Ref:19/SC/0520).

**Planned outputs:** OX-CHRONIC will provide detailed data on the evolving cognitive profiles of stroke survivors over several years post-stroke. Estimates of long-term prevalence as well as the effect of changes in cognitive profiles on mood, fatigue and quality of life will be examined. This Study is funded by a Priority Programme Grant from the Stroke Association (SA PPA 18/100032).

## INTRODUCTION AND RATIONALE

Stroke is the second leading cause of death and the third most important cause of disability burden in the world^1 2^. There are 1.2 million stroke survivors in the UK, with 100 000 new strokes every year^3^. Whilst mortality rates for stroke have decreased, principally due to better acute stroke care^4^, this improved stroke survival has resulted in a higher prevalence of chronic stroke survivors^5^.

Whilst Stroke can result in long-lasting physical^6^ and communication^7^ impairments, many of the less visible consequences are changes in affected mood^8^, wide ranging cognitive impairments^9–12^ and fatigue.^13^ Post-stroke cognitive impairment longer term is extremely common: in the largest ever national survey conducted with over 11,000 stroke survivors, 90% of respondents reported experiencing cognitive problems.^14^ Whilst observed prevalence data vary depending on the nature of the measures and the included cohort^10 12^, almost all stroke survivors suffer at least one cognitive domain deficit^9 11^ in the early stages post-stroke. Prevalence of cognitive impairment one year post-stroke has been estimated at 34%^15 16^. Cognitive impairments post-stroke negatively affect the level of social participation,^17^ mood,^18^ and quality of life,^19^ over and above the physical impairments and level of disability. Furthermore, a recent systematic review of quantitative studies on the unmet care needs of community-dwelling stroke survivors reported that the most frequently reported unmet needs were in the area of cognition (41.92%), followed by mood (40.13%).^20^

Post-stroke cognitive impairment is complex. It is currently comprised under the umbrella term of vascular cognitive impairment, defined as impaired cognitive function surpassing what is considered ‘normal’ within the ageing process, occurring in the presence of underlying vascular pathology.^21^ However, in predicting the natural history of post-stroke cognition, it is important to distinguish between ‘domain specific’ and ‘domain-general’ problems^22^. The NICE guidelines for post-stroke cognitive assessment,^23^ explicitly state to “as soon as possible … assess different cognitive domains post-stroke: attention, memory, spatial awareness, apraxia, and perception”. Additionally, the RCP clinical guideline for stroke^24^ states that each cognitive domain should not be considered in isolation. The Oxford Cognitive Screen (OCS) was designed with these distinct requirements in mind, as it is a domain-specific cognitive screen designed specifically for stroke patients ^9 25^. Its reach and significance as the routine first-line cognitive screening tool in stroke has been steadily increasing. The OCS has been widely adopted for clinical use within the NHS, as well as various sites internationally, as it has been culturally and language adapted in 7 other countries (e.g. ^26–28^) with more currently under development.

Studies concerning detailed domain-specific cognitive impairments initially post-stroke generally demonstrate some improvement over time in specific cognitive domains, such as spatial inattention/neglect,^29^ aphasia,^30^ and apraxia ^31^. However, data from the OCS screening study measuring individual domain-specific impairments show significant proportions of lasting impairment^32^ and similarly, a study investigating cognitive impairment post-stroke using a global cognitive screen found an overall prevalence of 56% at 6 months.^33^

Additionally, stroke patients are at an increased risk of developing dementia, as a meta-analysis reported 10% develop dementia after first-ever stroke and 33% after recurrent stroke^34^. However, in stroke survivors who do not progress to dementia diagnoses, prevalent yet often more subtle impairments are frequently overlooked, perhaps due to some individuals maintaining a degree of personal independence despite poor cognitive recovery. To better understand the nature and trajectories of long-term post-stroke cognitive impairment, we must delineate cognitive profiles on a fine-grained neuropsychological level and investigate their relation to long-term outcomes.

OX-CHRONIC is a prospective cohort study of stroke survivors who were initially recruited via consecutive sampling in an acute stroke ward and agreed to further assessments during long-term recovery.

## METHODS AND DESIGN

### Study aims and objectives

OX-CHRONIC capitalises on the extensive cognitive screening work in the Oxford Cognitive Screening project. The Primary aim of this study is to determine the prevalence and nature of cognitive impairments during long-term recovery from stroke (over 2 years post-stroke). There are four Work packages (WPs); WP1 covers the primary aim to provide in-depth neuropsychological profiling of domain-specific interrelations and trajectories in long-term stroke recovery. The primary objective here is

1. WP1: Identifying the prevalence of clinical impairment in six specific cognitive domains (language abilities, number processing, apraxia, attention, episodic memory, and executive function) in long-term stroke survivors using the Oxford Cognitive Screen (OCS).

Secondary objectives include:

1. WP1: Examining the impact of acute domain specific impairments on long-term cognitive profile trajectories.
2. WP2: Identify neuropsychological predictors of post-stroke dementia.
3. WP3: Identify the cognitive underpinnings of post-stroke mood disorders and fatigue, and the longitudinal relationships between PSCI and mood disorders and fatigue.
4. WP3: Identify the longitudinal relationships between PSCI and long-term outcomes in quality of life.

These study objectives were set out in the original funding application. The OX-CHRONIC dataset will continue to exist as a resource for future research use.

## PATIENT POPULATION - INCLUSION AND EXCLUSION CRITERIA

OX-CHRONIC directly leverages existing data. A cognitive screening programme based within the acute stroke unit, capturing the Oxfordshire area has been ongoing since 2012. Participants in this study have been assessed for stroke specific cognitive impairments during acute recovery and at six-month follow-up.^9 25^ The only exclusion criteria are an inability to stay alert for 20 mins and the lack of English language. Participants agree to be followed up 6 months later with a home visit to assess psychological consequences of stroke and quality of life. Participants who completed the follow up and gave explicit opt-in consent for re-contacting for further research will be contacted to take part in this two-year follow-up study (see Figure 1 for details regarding patient recruitment and attrition). Importantly, there is a minimum two-year interval between stroke and the first new assessment for OX-CHRONIC. The study is conducted according to the Declaration of Helsinki and approved by the local ethics committee (REC Ref:19/SC/0520). All participants provide informed or consultee consent before being included in the study.

**Figure 1.**
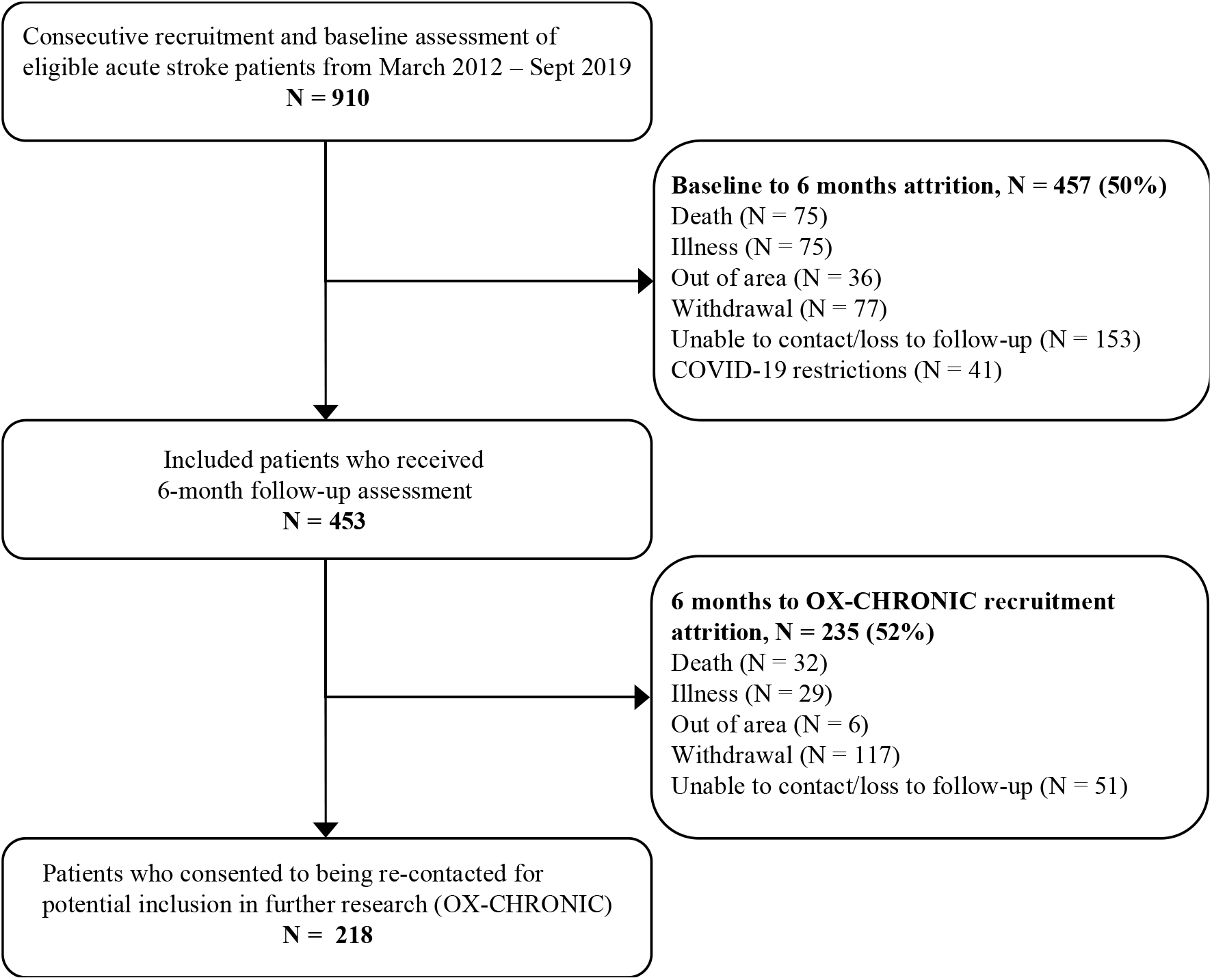
Flow-chart of participant recruitment and attrition.

Recruiting through our previous study means that we will be able to include participants who might otherwise not put themselves forward for research, including those with communication difficulties, but also those who feel depressed and/or isolated. We estimate we will be able to recruit at least 120 participants, accounting for attrition, to the first wave of the longitudinal cohort study. We estimate further attrition to the second timepoint of up to 20%.

Given the initial acute cohort of 910 stroke survivors, and the attrition which has already occurred (Figure 1), we have estimated the total completed cohort of this main study to consist of 96 participants. In addition to stroke survivors, we will also recruit carer participants to report on their experience of caring for long-term stroke survivors. Inclusion and exclusion criteria are shown in Table 1.

**Table 1.** Inclusion/exclusion criteria for stroke survivors and carers.

| Stroke Survivor Participants | Carer Participants |
| --- | --- |
| Inclusion Criteria |  |
| • Patient is willing and able to give informed consent OR positive opinion from a consultee is provided by a family member or carer<br>• Previous cognitive assessment with the following data from the two previous assessment time points:<br>○ Acute assessment (within 4 weeks of stroke):<br>▪ Neuroimaging – CT or MRI; lesion aetiology and lateralization<br>▪ National Institutes of Health Stroke Scale (NIHSS)<br>▪ Oxford Cognitive Screen (OCS)<br>○ Follow-up assessment (6 months) completed with OCS and broader daily functioning, Barthel, Stroke Impact Scale (SIS) etc. | • Adults aged 18 years or above<br>• Carer of a long-term stroke survivor<br>• Self-reports to know the stroke survivor well |
| Exclusion Criteria |  |
| • The patient is too unwell to be able to stay awake or concentrate for 20 minutes<br>• The patient or consultee/proxy has insufficient English comprehension to complete assessments, | • Lacks ability to give informed consent |

### Existing cohort information

Table 2 summarises the standardized measures used to collect data at each stage of the longitudinal study. Further demographics, any other neurological events since acute assessment, basic motor ability measures will be collected at each visit as routine. The Caregiver Strain Index^35^, Informant Questionnaire on Cognitive Decline in the Elderly^36^, and the Informant Assessment of the Geriatric Depression Scale^37^ will be gathered if there are willing carers present.

**Table 2.** Assessment tools used in the long-term waves of OX-CHRONIC

| Focus | Assessment Tool | Study Phase |  |  |  |
| --- | --- | --- | --- | --- | --- |
|  |  | Acute Assessment | Six-Month Follow-up | OX-CHRONIC Wave 1 | OX-CHRONIC Wave 2 |
| General cognitive status | Montreal Cognitive Screen<sup>43</sup> |  |  | X | X |
| Cognitive impairment profile | Oxford Cognitive Screen<sup>9</sup> | X | X | X | X |
| Language | Boston Naming Test (short)<sup>44</sup> |  |  | X | X |
| Verbal Fluency | Letter Fluency<sup>45</sup> |  |  | X | X |
|  | Category Fluency<sup>46</sup> |  |  | X | X |
| Executive Function | Hayling Test<sup>47</sup> |  |  | X | X |
|  | Trails Making Test A and B<sup>48</sup> |  |  | X | X |
|  | OCS-Plus Trails<sup>49</sup> |  |  | X | X |
| Working Memory | Digit Span forwards and backwards<sup>50</sup> |  |  | X | X |
| Episodic Memory | Logical Memory Test Part 1 & 2 |  |  | X | X |
|  | Picture Memory Test<sup>50</sup> |  |  | X | X |
| Visual Spatial Ability | OCS-Plus Figure Copy<sup>49</sup> |  |  | X | X |
|  | Rey Complex Figure Copy<sup>51</sup> |  |  | X | X |
| Spatial Attention | Star Cancellation test<sup>49</sup> |  |  | X | X |
| Language Discourse | Cookie Theft Picture Description Task<sup>52</sup> |  |  | X | X |
| Mood | Hospital Anxiety and Depression Scale<sup>53</sup> |  | X | X | X |
|  | Geriatric Depression Scale<sup>54</sup> |  |  | X | X |
|  | Apathy Evaluation Scale<sup>55</sup> |  |  | X | X |
| Fatigue | Fatigue Severity Scale<sup>56</sup> |  |  | X | X |
|  | Sleep Condition Indicator<sup>57</sup> |  |  | X | X |
| Cognitive Status | Cognitive Reserve Index |  |  | X | X |
|  | Cognitive Failures Questionnaire<sup>55</sup> |  | X | X | X |
| Quality of Life | Stroke Impact Scale<sup>58</sup> |  | X | X | X |
|  | EQ-5D-5L<sup>59</sup> |  |  | X | X |
| Activities of daily living and disability | Barthel (short form)<sup>60</sup> | X | X | X | X |
|  | Nottingham Extended ADL<sup>61</sup> |  |  | X | X |
|  | Modified Rankin Scale<sup>62</sup> |  |  | X | X |
| Care-giver questionnaires | Informant Questionnaire on Cognitive Decline in the Elderly (QCODE)<sup>36</sup> |  |  | X | X |
|  | Caregiver Strain Index<sup>35</sup> |  |  | X | X |
|  | Informant Assessed Geriatric Depression Scale<sup>37</sup> |  |  | X | X |

### Assessments during the OX-CHRONIC follow-up

First assessments started in February 2020, however, due to COVID-19 related disruptions, recruitment was suspended in March 2020 and could only restart in December 2020, following a major amendment of the study protocol to pivot to remote testing via telephone or video call.

The purpose of the long-term follow-up assessments is to provide in-depth neuropsychological profiling using clinical screening tools in combination with more detailed cognitive testing batteries. Tests and questionnaires will be identical between both waves. Assessment tools were selected based on extensive use in research with stroke survivors in which acceptable levels of reliability and validity have been reported. Furthermore, the overall package of assessment tools was designed to limit the burden on participants. We achieved this by piloting assessments with stroke survivor representatives from our Study Management Team and Steering Committee. As a result, the test battery should provide in-depth assessments that will accurately capture the psychological consequences of stroke in a way that the results can be related back to existing literature. Alongside assessment results, detailed records of problems interfering with testing will be taken (e.g. poor vision, functional issues of dominant arm, fatigue). Such information will be used for subjects with incomplete testing when assessing their study records for diagnosis of impairment and DSM-V criteria for dementia. This will help to avoid spurious classifications of cognitive impairment and dementia.^38^

Due to COVID-19 disruptions, participants will be assessed remotely by trained research assistants with backgrounds in Speech and Language therapy, psychology or occupational therapy either over telephone or video conference. While there are other studies that use remote testing to monitor cognitive change after stroke (e.g.^39^), the in-depth neuropsychological testing in OX-CHRONIC has not all been designed or validated for remote assessments, though further validation in an independent cohort is planned. Testing packs with materials will be sent out and trained assessors will conduct the tests through a series of calls up to a duration of 1-hour each. The cognitive tests and questionnaires are indicated in Table 2. In addition, participants will be given an option to wear an activity tracker for a one-week period and levels of activity will be recorded.

The switch from in-person to remote testing required some adjustments to how tests were administered. For example, the MoCA has been adapted for telephone administration^40^ but here we will use the whole of the MoCA, with visual stimuli and in-put sections sent in the post to be completed by participants. In the event that these sections prove not to be valid via remote testing, we will still have the portion of the MoCA that has been established as reliable over the phone. A similar approach will be taken with other tests, where visual stimuli or input fields are required, they will be sent via post and the participant will be instructed on when to open and complete testing packs by the assessors over the phone. Then, during an assessment call, the trained assessor will talk each participant through the packs, instructing them how to fill out forms or asking for verbal (or non-verbal e.g. tapping phone) responses to tasks. In consultation with stroke survivor representatives, it became clear that the most burdensome part of our planned assessment would be filling out long questionnaires – this is partly due to it being less engaging than cognitive assessments. Therefore, where possible, we switched to short form questionnaires and assessments e.g. the SIS^41^ and Barthel^42^. After all assessments were complete, the participants will put the packs back in prepaid envelopes and mail them back.

In addition to these assessments, relevant baseline clinical data pertaining to the participant’s stroke admission will be extracted from electronic medical records, such as existing comorbidities (e.g. delirium, known pre-existing cognitive problems, atrial fibrillation, hypertension, diabetes mellitus, smoking habits, previous stroke), discharge information (date, destination, cognitive status, length of stay, mortality), and long term outcomes such as final diagnoses and death.

### PRIMARY OUTCOMES

The primary outcome for OX-CHRONIC will be the cognitive profiles of domain-specific impairments and how they progress during long-term recovery. This will include the percentage of participants who have domain specific impairments as defined using normative cut-off scores on the OCS assessments as outlined in Demeyere et al, (2015)^9^. Furthermore, dementia status using DSM-5 criteria and based on medical records, cognitive testing and impairment in function of activities of daily living will be assessed. This will include existing dementia diagnoses explicitly referred to in medical records as well as evidence of meeting DSM-5 criteria based on medical records or standardized testing carried out in the study and assessed by two trained physicians, see Pendlebury et al, 2015^38^ for further details). This approach of using medical records in combination with remote testing will help mitigate attrition. For example, if individuals are not able to complete OX-CHRONIC testing but their medical records include end point information such as dementia, discharge destination, or death, this can be used in proportional hazard modelling.

### SECONDARY OUTCOMES

Secondary outcomes will include other psychological consequences of stroke such as mood disorders and fatigue. Additionally, we will collect data on cognitive status, quality of life, and function of activities of daily living and disability. All assessments are listed in Table 2 and standardized cut-offs will be used where appropriate.

### DATA MANAGEMENT

Data storage will be facilitated using Research Electronic Data Capture (REDCap). REDCap provides a secure storage solution with easy-to use forms with real-time field validation. The University of Oxford IT service provides support and monitoring for the REDCap application. All data will be de-identified before being stored on REDCap.

## STATISTICAL ANALYSES AND POWER CALCULATIONS

Statistical analyses for the primary outcomes will include time to event analyses such as Cox proportional hazard models and linear mixed effects models. Cox proportional hazard models will be used to investigate the association between acute cognitive profiling and long-term outcomes such as persistent PSCI or dementia. For assessing the trajectories of psychological profiles or other important outcomes, such as quality of life over time, linear mixed effect models will be employed (e.g., to assess the difference in the rate of increased depressive symptomology between participants with and without acute cognitive impairments). All models will include known covariates based on the literature (e.g.^38^) and statistical screening between outcome groups.

The *powerEpiCont* function from the R package *powerSurvEpi*^*63*^ was used to calculate power for Cox regression predicting final cognitive impairment or dementia status with a prevalence of 33%^16^, using acute OCS raw score mean and standard deviations. Table 3 shows that while several of OCS subtests reach sufficient power to detect a hazard ratio (HR) of 1.25, most are only suitably powered to detect HR of 1.5 or greater. However, based on previous research, it is expected that HR values will range between 3-4.

**Table 3.**
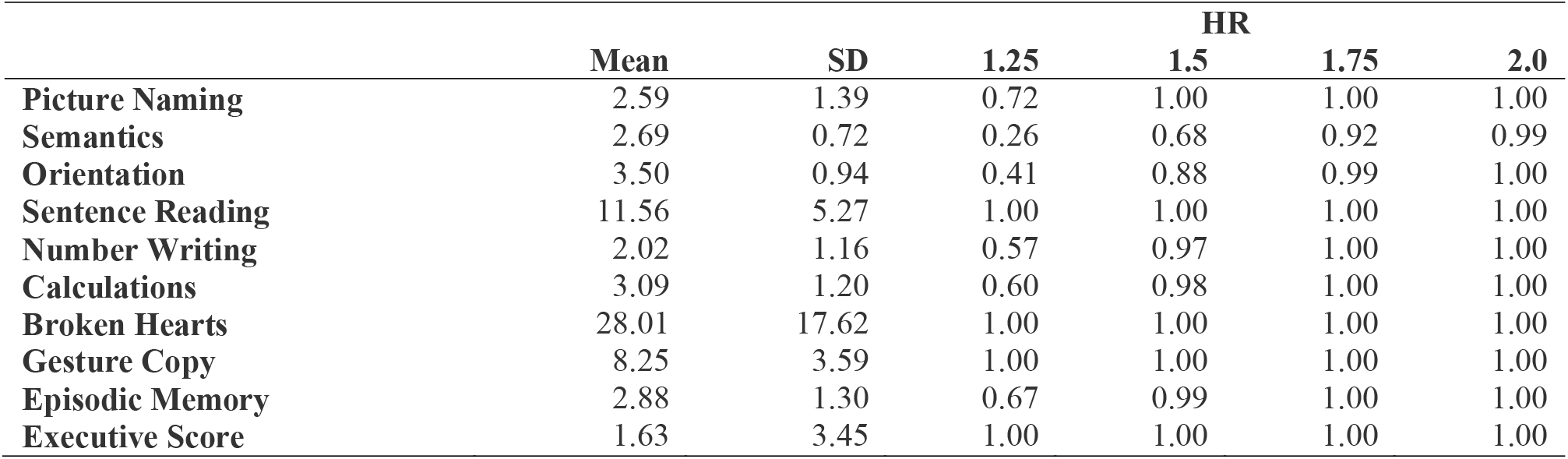
Estimated power to detect risk of persistent cognitive impairment or dementia status at 33% prevalence using the mean and standard deviation for individual OCS test scores, shown for different postulated hazard ratios

The *powerSim* function from R package *simr*^*64*^ was used to estimate power for linear mixed effects models to detect a different in the average rate of change in a dependent variable (e.g. depression score) between individuals with and without an acute cognitive impairment at a 30% prevalence rate of cognitive impairment. Assuming four timepoints of depression data, for 96 participants and a medium effect size of 0.5 for difference in mean slopes, the power will be 1.0 (95% confidence intervals: 99.63 - 100.0).

## STUDY ORGANIZATION AND FUNDING

OX-CHRONIC Study Management Team constitutes co-investigators and two stroke survivor representatives. The Study Management Team meets every six months, in person or via videocall. A Study Steering Committee also offers oversight. This Committee includes the study representative, three external and independent academics (expertise in Psychology, Occupational Therapy and Speech and Language Therapy), a stroke survivor representative, and a representative of the funder. A senior academic, Prof Audrey Bowen, chairs the committee. The Steering Committee also meets every six months to provide oversight, and strategic direction. The work is organized in work packages (as outlined in the study aims section) overseen by different members of the Management Team.

OX-CHRONIC is funded by a Priority Programme Grant from the Stroke Association (# SA PPA 18/100032)

## DISCUSSION

We will investigate the co-occurrences and time-course of recovery from acute initial assessments to longer term (>2 years) life after stroke for the 5 specific cognitive domains found to be impaired at high frequencies in the acute stage: Language abilities (including naming and reading); Number abilities; Apraxia; Attention (controlled attention and spatial attention – e.g. hemi-spatial neglect (both object and space based) and Episodic Memory (verbal and visual).

Using these stroke specific cognitive profiles, we will investigate whether the presence of domain-general cognitive deficits impedes recovery of domain-specific cognitive impairments, as well as identify participants who are developing a degenerative dementia. Importantly, the extensive neuropsychology will help differentiate between different degenerating cognitive domains.

With respect to other psychological consequences of stroke, though cognitive impairment has been consistently associated with depression and anxiety^18 65^, the temporal nature of the relationship, and which cognitive domains may be most associated with more severe mood disorders remain unclear. We will use in-depth cognitive profiling and mood screening to assess these relationships over the long-term recovery of stroke patients. Fatigue has been reported to be one of the most distressing symptoms after stroke^66^ and was highlighted by a large survey in stroke survivors as an unmet need^67^. We will investigate the temporal relationship between cognitive profiles of impairment and fatigue. Furthermore, by leveraging activity monitoring technology, we will evaluate the relationships between post-stroke fatigue and activity levels. This will allow us to examine the factors that may differentiate between those with post-stroke fatigue who manage to be active and those who do not.

A key strength of this study is the assembly of a rich clinical dataset including demographic, extensive clinical data (comorbidities, stroke characteristics, frailty, stroke complications, delirium), clinical brain imaging etc. along with in-depth cognitive profiling. This will allow us to determine the independent predictive value of domain specific and domain general cognitive dysfunction in longer-term outcomes while controlling for other important factors. Furthermore, the combination of follow-up testing with data extraction from medical records will mitigate the effect of attrition on data analyses. This will have the overall effect of protecting against underestimation of dementia diagnoses.^38^

### THE IMPACT OF COVID-19 ON THE STUDY DESIGN

The COVID-19 pandemic presented an unforeseen challenge in recruiting and assessing stroke participants for the OX-CHRONIC study. Originally designed to include home-visits to complete in-depth neuropsychological assessments, the protocol had to be revised to guarantee the study could proceed while minimizing the risk of infection in a clinically vulnerable population. When it became clear that COVID was going to be a long-term threat, the OX-CHRONIC study team decided to switch to remote testing for both waves of the long-term follow-up. This approach had the advantage of being resilient to any further disruptions due to national or local lockdowns. However, it did present some challenges. Several assessments were not suitable for remote testing while all others were not designed nor evaluated for use over telephone or video call. We settled on a method using paper-based assessments that will be mailed to participants along with some assessments that can be completed by verbally responding over the phone. This hybrid method of paper assessments being applied by a remote assessor will require validation. Therefore, an unexpected work package for the OX-CHRONIC project will be to validate the remote testing packs against in-person testing. Due to the logistics and burden of multiple testing sessions, this validation work will be carried out on a separate group of stroke survivors, independent of OX-CHRONIC participants.

## SUMMARY AND CONCLUSIONS

Currently, clinical stroke services do not routinely follow up patients, and do not provide long term input to the primary care teams caring for long term stroke survivors. With regards to psychological consequences, the knowledge gap is even greater. Cognition and mood are not routinely assessed after the acute admission, and clinical stroke services rarely engage with specialist psychology services. The specific long-term information gained from this study will help inform how services are delivered. This research will help inform specific guidelines on how to use initial cognitive assessment data to plan future care, how often stroke survivors should be assessed and what these assessments should look like. Within the primary care services, understanding stroke specific residual cognitive problems and broader psychological consequences such as fatigue and mood changes will help manage patients and engage with community psychology services.

## Data Availability

Data collection is ongoing and will not be available at this time.

## Notes

### Competing Interest Statement

The authors have declared no competing interest.

### Clinical Trial

ISRCTN81038194

### Author Declarations

This study was approved by South Central - Berkshire Research Ethics Committee, Bristol REC Centre, Whitefriars, Level 3, Block B, Lewins Mead, Bristol, BS1 2NT. (REC Ref:19/SC/0520)

